# Grey Matter Iron and Neuromelanin in Psychosis: A Systematic Review and Meta-Analysis of MRI Studies

**DOI:** 10.64898/2026.01.15.26344182

**Authors:** Luke James Vano, Jan Sedlacik, Richard Włodzimierz Carr, Bernard Roch Bukala, Oliver David Howes, Robert Ali McCutcheon

## Abstract

**Objective:** The pathophysiology of psychosis remains unclear. Preclinical, postmortem, and imaging evidence implicates iron and neuromelanin, but the consistency and magnitude of effects are uncertain. We aimed to characterise brain iron and neuromelanin alterations in psychosis through a systematic review and meta-analysis of iron-sensitive MRI and neuromelanin-sensitive MRI (NM-MRI) studies.

**Methods:** We searched EMBASE, PubMed, and PsycINFO from inception to October 31, 2025, for case–control studies using iron-sensitive MRI or NM-MRI in patients with psychosis. We used random-effects models to calculate effect sizes (Hedges g) and meta-regressions to examine clinical confounders. The primary outcomes included effect sizes for NM-MRI and iron-sensitive MRI measures—transverse relaxation rate (R2), effective relaxation rate (R2*), and quantitative susceptibility mapping (QSM).

**Results:** Twenty-seven reports, including 879 individuals with psychosis and 813 controls, were analysed. Meta-analyses were conducted across the caudate nucleus, putamen, globus pallidus, thalamus, and substantia nigra. In psychosis, R2* was significantly lower across all examined regions (g= –0.27 to –0.40), QSM values were lower in the substantia nigra (g= – 0.61; 95% CI, –0.84 to –0.38), and R2 was lower in the caudate nucleus (g= –0.30; 95% CI, – 0.56 to –0.04). NM-MRI values in the substantia nigra were significantly higher (g= 0.39; 95% CI, 0.23 to 0.55), though this effect strongly correlated with chlorpromazine daily equivalent dose (β= 0.001; 95% CI, 0.0003 to 0.0018), suggesting medication-related effects.

**Conclusions:** Psychosis is associated with lower subcortical iron-sensitive MRI values. This was most marked in the substantia nigra, where NM-MRI values—which index neuromelanin-bound iron in dopamine neurones—were significantly higher. This suggests that while subcortical iron is overall lower in psychosis, neuromelanin-bound iron is increased within dopamine neurones. Investigating the mechanisms underlying iron alterations may provide new treatment targets.

## Introduction

Psychotic illnesses, including schizophrenia and bipolar affective disorder, are leading causes of global disease burden^1^. Dysregulation of dopamine synthesis^2^, myelin formation^3^, and mitochondrial respiration^4^ have all been implicated in the pathophysiology of psychosis. As iron is an essential cofactor for all three processes^5^, altered brain iron may play a key role in disease mechanisms.

Epidemiological studies link maternal iron deficiency to a higher risk of psychotic disorders in offspring^6^. Additionally, low blood iron is associated with a diagnosis of schizophrenia^7^ and negative symptoms^8^. Postmortem studies assessing brain iron in schizophrenia have produced inconsistent results^9–11^. As these may reflect confounders specific to postmortem analyses (e.g., differences in the mode of death), in vivo assessment with iron-sensitive MRI may help clarify the magnitude and nature of brain iron alterations in psychosis.

Most iron in dopamine neurones is neuromelanin-bound^5^. Neuromelanin forms through iron-dependent oxidation of cytosolic dopamine and is not readily degraded^5^, meaning it reflects longer-term dopaminergic activity. Neuromelanin-sensitive MRI (NM-MRI) detects the neuromelanin-iron complex, but is insensitive to either iron or neuromelanin alone^12^. Because iron-sensitive MRI indexes all forms of iron^13^, comparing NM-MRI with iron-sensitive MRI in the substantia nigra can help clarify the biological origin of iron abnormalities in psychosis.

Brain iron in psychosis has been assessed using MRI measures that increase with iron concentration, including the transverse relaxation rate (R2), the effective transverse relaxation rate (R2*), and quantitative susceptibility mapping (QSM)-derived magnetic susceptibility (χ). Yet no meta-analysis has yet synthesised these findings, leaving the magnitude and consistency of case–control differences uncertain. Since previous NM-MRI meta-analyses^14,15^, the sample size in the psychosis group has more than doubled, warranting an updated analysis. To address these gaps, we perform a meta-analysis of iron-sensitive MRI and NM-MRI studies in psychosis.

## Methods

We followed the Preferred Reporting Items for Systematic Reviews and Meta-Analyses (PRISMA) guidelines for this review^16^. We registered our protocol on PROSPERO (https://www.crd.york.ac.uk/PROSPERO/view/CRD420250637198). The full study selection strategy is detailed in the Supplementary Methods.

### Study selection

We searched EMBASE, Pubmed, and PsycINFO databases, from inception to 31^st^ October 2025, using the following base terms: MRI AND (QSM OR iron OR susceptibility weighted imaging OR relaxometry OR gradient recall echo OR T2 OR R2 OR R2* OR T2* OR neuromelanin) AND (psychosis OR schizo* OR psychotic OR bipolar disorder OR manic OR bipo*). All studies reporting R2, R2*, QSM, or NM-MRI for any grey matter regions-of-interest (ROIs) in patients with psychosis in comparison with healthy controls were included.

## Meta-analysis

### Data collection

We extracted the mean and standard deviation (SD) of the R2, R2*, χ, and NM-MRI values for the psychosis and control groups. When the transverse relaxation time (T2) was reported, this was converted to R2. When separate left and right ROI statistics were produced, bilateral values were calculated by weighting values according to volume; otherwise, equal weighting was applied.

The following potentially moderating factors were also recorded: age, sex, psychiatric diagnosis, antipsychotic medication chlorpromazine daily equivalent (CPZE) dose, duration of illness, and Positive and Negative Symptom Score (PANSS) subscores. To enable cross-study comparisons, we converted antipsychotic doses to CPZE dose and symptom scores to PANSS subscores via methods detailed in our Supplemental Methods. Study quality was assessed using the Newcastle–Ottawa Scale, with the exposure category removed given its irrelevance for imaging studies^17^.

### Mean differences

Statistical analyses were conducted when four or more reports examined an ROI with a specific MRI modality; otherwise, study results were discussed in the narrative review. For statistical analyses, overall effect size (Hedge’s g) and 95% confidence interval (CI) were calculated using a random-effects model (p<0.05) to account for expected between-study variability. Analyses were performed in R (v4.5.0)^18^ using “metafor” (v4.8.0)^19^.

### Sensitivity analysis

Robustness of the effect size was evaluated using the leave-one-out method. Inconsistency between studies was examined using the I^2^ statistic, with significance assessed with Cochran’s Q test (QEp<0.05)^20^. When heterogeneity was present, outliers were identified when Cook’s distance was >4/number of studies^21^. An additional analysis was performed with these excluded to assess their impact on the pooled effect size and heterogeneity estimates.

### Variability analysis

Group differences in variability were assessed using the log SD ratio. Significant log SD ratios were meta-regressed on the pooled mean to determine whether variability differences reflected mean scaling

### Meta-regression with clinical covariates

We used meta-regression in our mixed-effects model to evaluate whether the following confounders influenced the outcomes of the meta-analysis: average age of study participants, proportion who were male, average CPZE dose, duration of illness, chronic illness (study-defined or when mean duration of illness >10 years), PANSS total, positive, and negative subscores. If significant confounders were categorical, meta-analysis was performed by subgroup.

### Assessment of bias

When ≥10 reports were available for an analysis, we evaluated publication bias by visually inspecting forest plots and using Egger’s regression test for small-study effects^22^.

### Narrative review

A narrative review of ROIs examined by fewer than four reports, voxelwise analyses, and the impact of specific psychotic diagnoses on effect size is provided in the Supplemental Materials.

## Results

### Study selection

Of the 950 records identified, 27 were included in the review (Supplemental Figure S1). Three used both QSM and R2*^23–25^, three used R2* only^26–28^, three used QSM only^29–31^, six used R2^32–37^, and 12 used NM-MRI^38–49^. The included study characteristics are shown in Table 1 with a summary of their findings in Supplementary File 1. The mean Newcastle–Ottawa Quality Assessment Scale score of included studies was 4.92 (out of 6.0), indicating overall good methodological quality (Supplementary Table S1).

**Table 1.**
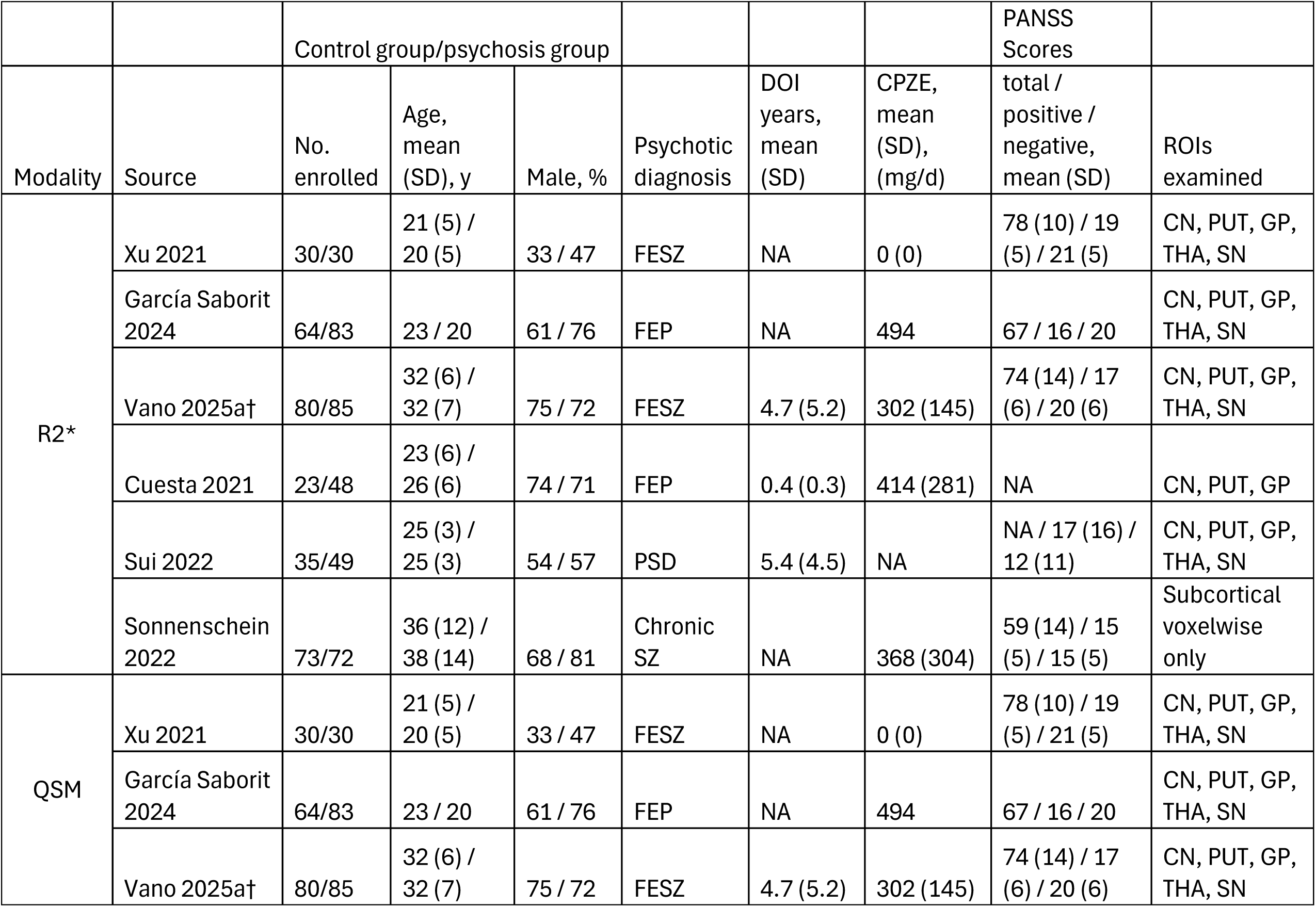
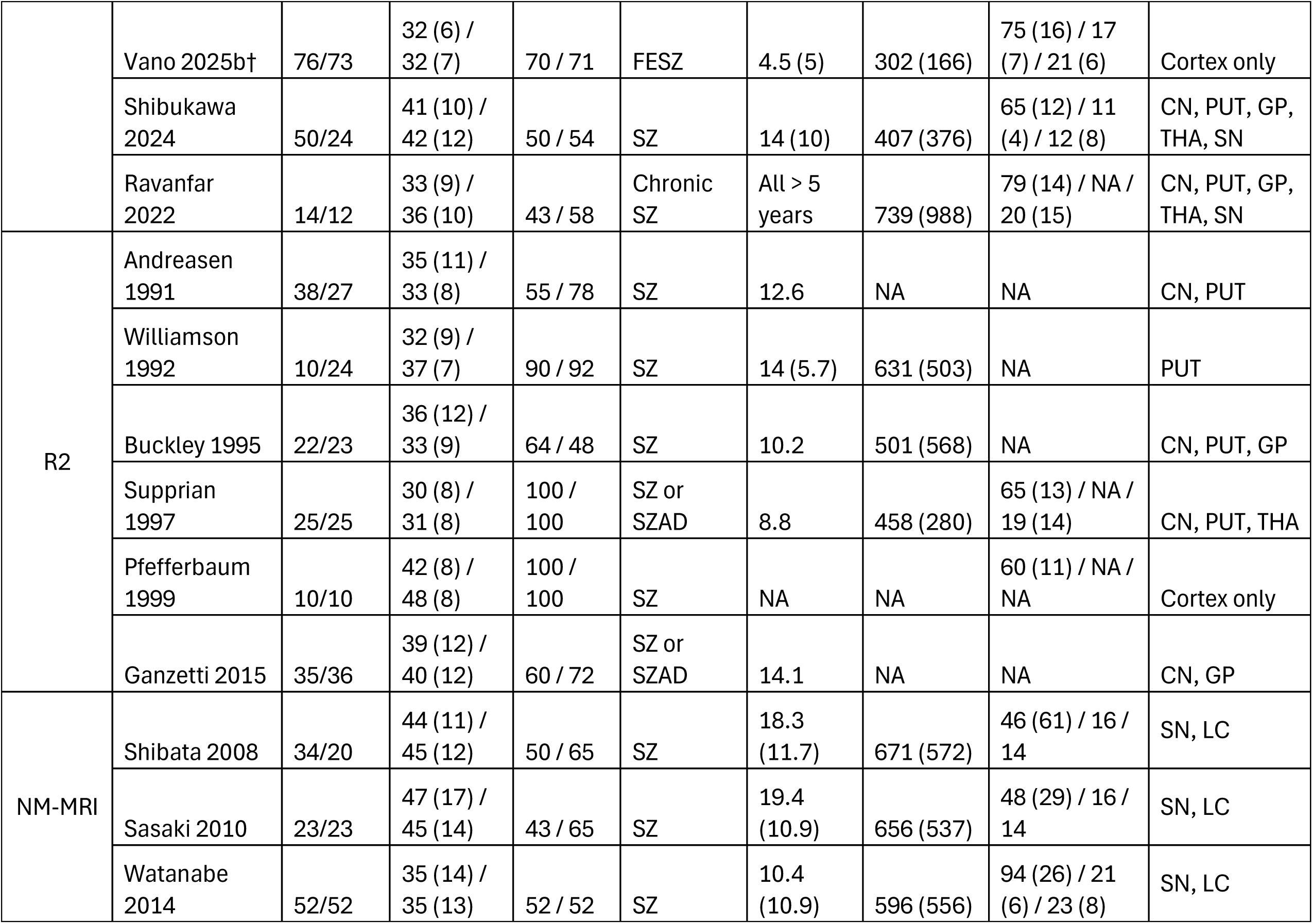
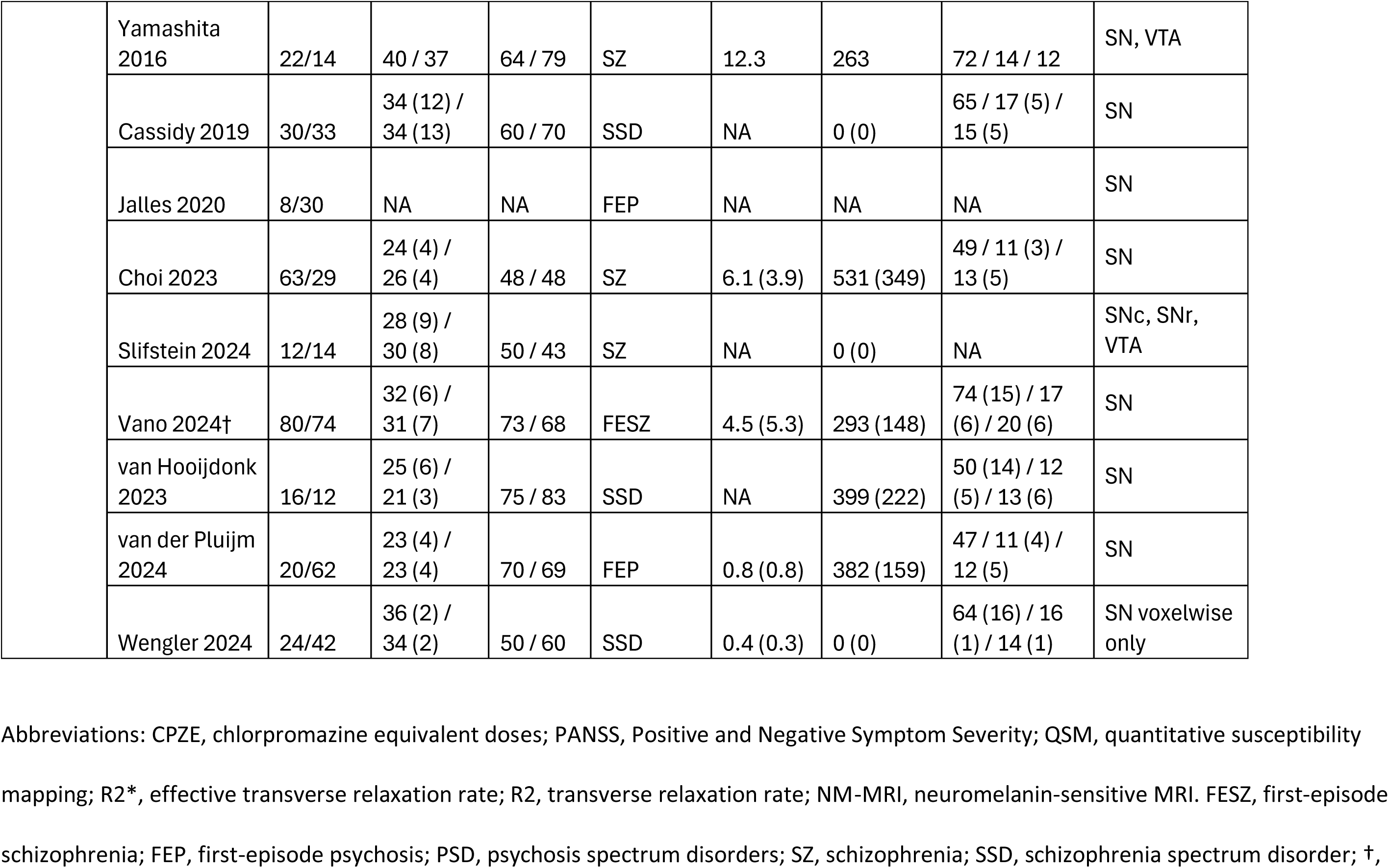
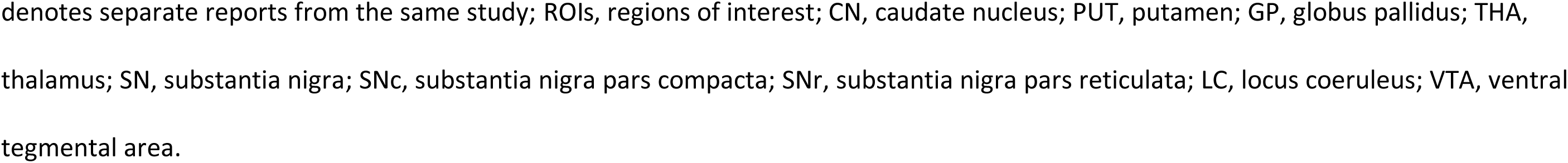
Meta-analysis study characteristics

### Mean differences

Summary effect sizes and heterogeneity estimates for the non-nigral ROIs are shown in Figure 1 and for the substantia nigra in Figure 2 (excluding an outlier QSM study). Study-level forest plots are presented in the Supplemental Materials (Supplementary Figures S2–S5).

**Figure 1.**
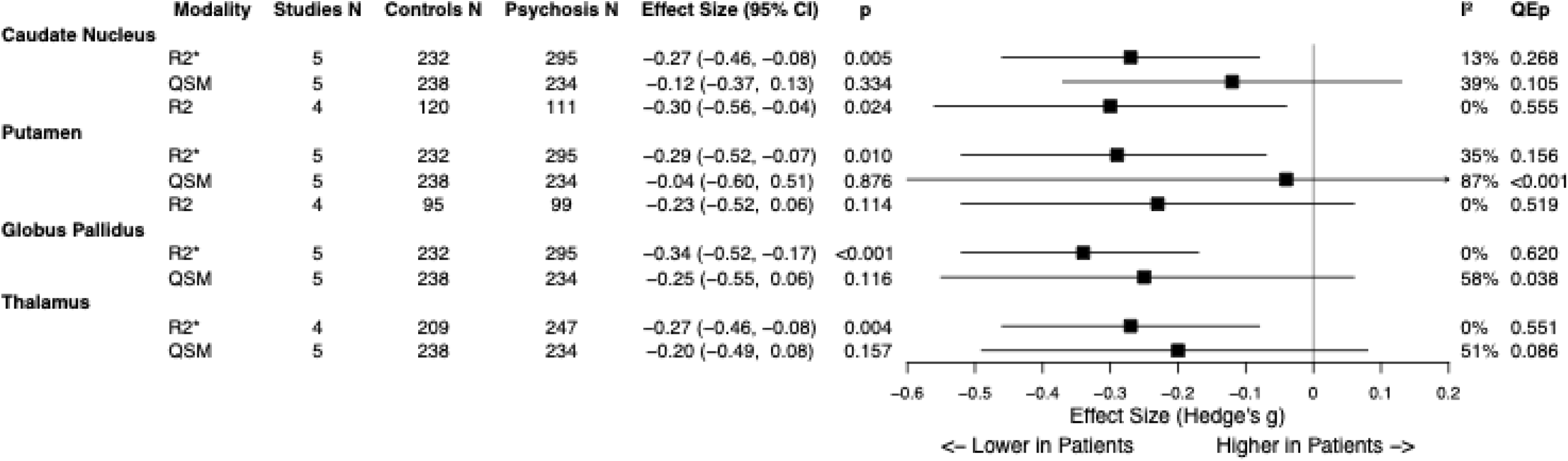
Summary plot showing effect sizes and heterogeneity estimates for each region of interest (ROI) and iron-sensitive MRI modality.

**Figure 2.**
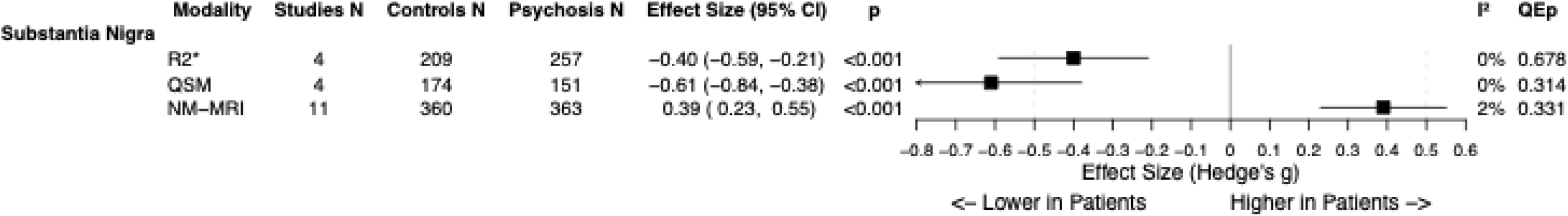
Summary plot showing effect sizes and heterogeneity estimates in the substantia nigra for the effective transverse relaxation rate (R2*), quantitative susceptibility mapping (QSM), and neuromelanin-sensitive MRI (NM-MRI).

Five R2* studies assessed the caudate nucleus, putamen, and globus pallidus, while four additionally examined the thalamus and substantia nigra. R2* was significantly lower in patients with psychosis than healthy controls across all ROIs: caudate nucleus (g= –0.27; 95% CI, –0.46 to –0.08; p=0.005), putamen (g= –0.29; 95% CI, –0.52 to –0.07; p=0.01), globus pallidus (g= –0.34; 95% CI, –0.52 to –0.17; p<0.001), thalamus (g= –0.27; 95% CI, –0.46 to –0.08; p=0.004), and substantia nigra (g= –0.4; 95% CI, –0.59 to –0.21; p<0.001).

While five QSM studies analysed the same ROIs noted above, χ was only significantly lower in patients with psychosis than controls in the substantia nigra (g= –0.44; 95% CI, –0.75 to – 0.12; p=0.007).

At least four R2 studies assessed the caudate nucleus and putamen, with the only significant case–control difference being lower caudate nucleus R2 in psychosis (g= –0.3, 95% CI, –0.56 to –0.04; p=0.024).

Across 11 NM-MRI studies, substantia nigra values were significantly higher in patients with psychosis than in controls (g= 0.39; 95% CI, 0.23 to 0.55; p<0.001).

### Sensitivity analysis

Significant heterogeneity was only detected for QSM analyses (Figure 1) of the substantia nigra (I²=61%, QEp=0.03), putamen (I²=87%, QEp<0.001), and globus pallidus (I²=58%, QEp=0.038). The only outlier result was χ in the substantia nigra for García Saborit et al (Cook’s distance=1.12, cut-off=1.00)^24^. χ values for both the control and psychosis groups in that study were substantially lower than in the four other datasets (Supplementary Figure S6). After removing this outlier, the effect size increased (g= –0.61; 95% CI, –0.84 to –0.38; p<0.001) and heterogeneity was abolished (I²=0%, QEp=0.314), suggesting a more robust analysis. Therefore, this result was eliminated from further analyses. All results remained stable under leave-one-out analysis (Supplementary Table S2).

### Variability analysis and meta-regression with clinical covariates

Results from the variability analyses are provided in Supplementary Table S2. Variability was only significant for R2 analysis of the putamen (Log SD ratio=0.30; 95% CI, 0.09 to 0.50; p=0.004). Meta-regression of the log SD ratio against the pooled mean was not significant (p=0.341), meaning that the greater variability in the psychosis cohort was not due to mean scaling.

### Meta-regression with clinical covariates

Results from the meta-regression are in Supplementary file 2. Higher CPZE doses were associated with greater NM-MRI effect sizes (k=10; β= 0.001; 95% CI, 0.0003 to 0.0018; p=0.006; Supplemental Figure S7). Higher PANSS total scores were associated with more negative χ effect sizes in the globus pallidus (k=5; β= –0.0525; 95% CI, –0.0955 to –0.0094; p=0.017).

First-episode illness was associated with larger negative χ effect sizes in the caudate nucleus (k=5; β= –0.5449; 95% CI, –1.0081 to –0.0817; p=0.021) and putamen (k=5; β= –0.9075; 95% CI, –1.4565 to –0.4846; p<0.001). Given this, we performed separate meta-analyses for the first-episode and chronic psychosis cohorts in these regions (Supplemental Figure S8).

In first-episode psychosis, χ was significantly lower in the caudate nucleus (g= –0.28; 95% CI, –0.49 to –0.08; p=0.008) and putamen (g= –0.45; 95% CI, –0.66 to –0.25; p<0.001) compared with controls, whereas no significant differences were observed in chronic illness (p=0.280 and p=0.136, respectively). R2* was also significantly lower in first-episode psychosis for the caudate nucleus (g= –0.31; 95% CI, –0.54 to –0.08; p=0.01) and putamen (g= –0.35, 95% CI, –0.58 to –0.12; p=0.004) compared with controls. There were no R2* studies in chronic illness. While there were no R2 studies in first-episode psychosis, R2 was significantly lower in chronic illness for the caudate nucleus (g= –0.38; 95% CI, –0.67 to – 0.08; p=0.013) and putamen (g= –0.36; 95% CI, –0.7 to –0.01; p=0.041).

### Publication bias

There were only enough studies to evaluate publication bias for the NM-MRI analysis, where inspection of the funnel plot (Supplementary Figure S9) and Egger’s regression test (p=0.304) indicated no evidence of bias.

## Interpretation

Our meta-analysis found lower subcortical iron-sensitive MRI values in participants with psychosis than in healthy controls. Specifically, R2* was lower across all five analysed subcortical ROIs, while R2 was lower in one of two ROIs. QSM-derived χ was only lower in the substantia nigra, the region with the largest R2* group difference. NM-MRI values in the substantia nigra—which index the neuromelanin-iron complex within dopamine neurones^12^—were elevated in psychosis. Taken together, this indicates that while total subcortical iron is lower in psychosis, nigral neuromelanin-bound iron within dopamine neurones may be increased.

Further insights may be gained by considering myelin, which increases R2^50^ and R2*^51^ but decreases χ^51^. Thus, the relatively consistent reductions in R2* and R2 and the inconsistent reduction in χ in psychosis may reflect concurrent loss of both iron and myelin, enhancing group differences in R2 and R2* but masking χ reductions due to iron loss alone. Figure 3 summarises these relationships and the meta-analytic results.

**Figure 3.**
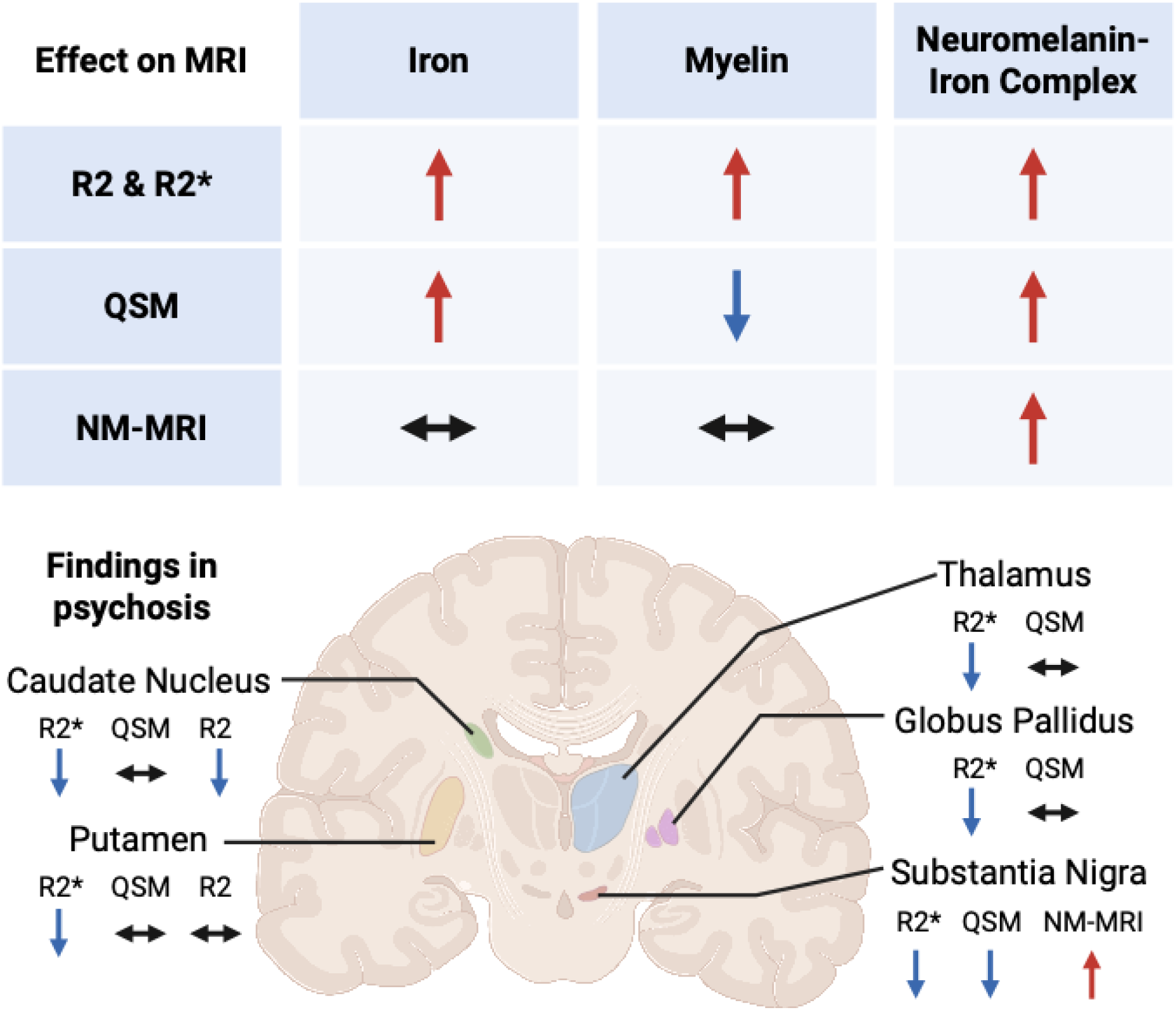
Comparison of iron, myelin, and neuromelanin effects on MRI modalities and results from the meta-analysis

### Strengths and limitations

Most R2* and QSM studies (>70% of participants) used both measures in the same cohorts^23–25^, suggesting that variation in R2* and χ effect sizes was not due to study differences. One dataset (controls N=80; psychosis N=74) had QSM, R2*, and NM-MRI^23,46^. In this sample, R2* and χ were lower for subcortical ROIs and NM-MRI values were higher in the substantia nigra for patients with psychosis, consistent with our overall meta-analytic results.

No individual studies correlating antipsychotic dose with R2*^23,25^, χ^23,25,29^, or NM-MRI^39,41,46^ have found a significant relationship. However, in our meta-analysis, CPZE dose was correlated with NM-MRI effect sizes. This suggests that greater antipsychotic use may contribute to elevated NM-MRI values in patients. One factor that may explain this association is age. The three studies with the highest CPZE doses had the oldest participants and longest illness durations^38–40^. As neuromelanin accumulates with age^5^, group differences may be greater in later life. Nevertheless, NM-MRI effect sizes did not correlate with illness duration or age, so we think this is unlikely to be driving the association. Another factor is that the studies with the highest CPZE doses delineated the substantia nigra more conservatively than the unmedicated studies^42,45^, which used larger, voxelwise-optimised ROIs. As previously discussed^52^, larger ROIs may dilute effects restricted to dopaminergic neurones.

Finally, it is possible that antipsychotic treatment leads to greater dopaminergic activity, and hence higher neuromelanin levels. However, a longitudinal study of dopamine synthesis capacity did not find evidence for dopaminergic changes in patients over a few weeks^53^.

Nevertheless, these may become apparent over the longer term. Longitudinal studies assessing the effect of antipsychotic medication on NM-MRI are warranted to resolve this uncertainty.

Only one R2* study directly compared different diagnostic categories of psychosis, finding R2* reductions for most ROIs in schizophrenia, reductions in half ROIs in schizoaffective disorder, and no differences in bipolar disorder^27^. While this could indicate that those with schizophrenia primarily drive effects, further research is warranted to assess this.

Significant heterogeneity was observed for QSM findings in the putamen, globus pallidus, and substantia nigra. Excluding one outlier (García Saborit et al^24^) resolved heterogeneity in the substantia nigra. In that dataset, χ values for the control group were markedly lower than in all other studies, with a notable proportion of negative values (Supplemental Figure S6). As no negative values were reported in a large multicentre cohort of 623 healthy adults^54^, this discrepancy suggests significant methodological differences from the included studies or data quality issues, supporting its removal.

Heterogeneity in the putamen was eliminated when separate analyses were completed for first-episode and chronic cohorts. First-episode psychosis was associated with significantly lower χ and R2* in the putamen and caudate nucleus, relative to controls. In the chronic cohort, R2 was lower in both ROIs, while χ was only numerically higher than controls. Iron accumulation with illness progression is unlikely to account for these findings, as it would be expected to increase R2. Instead, they may reflect greater myelin loss in chronic illness, which would both decrease R2 and increase χ. The small size of the chronic sample for the QSM analysis (N=36) necessitates testing in larger chronic cohorts.

Due to inconsistent cortical ROI labelling, we were unable to perform a meta-analysis of cortical ROIs. As discussed in our supplementary narrative review, three^32,33,36^ of six^32–37^ studies reported significantly lower R2 across various cortical ROIs. Conversely, the only QSM study reported significantly higher χ for the left temporo-parieto-occipital junction in psychosis without a difference in myelin-sensitive MRI^31^. The findings of this latter study are consistent with a recent, well-powered postmortem case-control study that reported cortical iron accumulation in patients with schizophrenia, which was most marked in younger individuals^11^. Because the QSM study was conducted in younger first-episode patients, whereas the R2 studies involved older populations with chronic illness, chronicity may account for these divergent results. Longitudinal iron-sensitive MRI studies following patients from illness onset are warranted to clarify the origin of cortical differences.

### The biological origin of altered iron-sensitive MRI and neuromelanin-sensitive MRI values in psychosis

Our interpretation that simultaneous iron and myelin loss in psychosis explains our iron-sensitive MRI findings is supported by our recent case–control study, which demonstrated lower subcortical χ in schizophrenia despite myelin-sensitive MRI techniques indicating that subcortical and white matter myelin were also lower^23^. In a similar case–control study, Sui et al. reported that lower R2* in psychosis for multiple subcortical ROIs was unrelated to differences in the macromolecular proton fraction, a marker of myelin^27^. Taken together, these data indicate that while iron loss is the primary driver of altered iron-sensitive MRI values in psychosis, concurrent myelin loss is likely an important confounder.

Oligodendrocyte loss may underlie our findings, given that they have the highest iron concentration of any brain cell^55^ and produce myelin. This is supported by postmortem studies reporting lower oligodendrocyte concentrations in the anterior putamen^56^, caudate nucleus^57^, and prefrontal cortex^58^, along with reduced brain expression of oligodendrocyte-related genes in patients with schizophrenia relative to controls^59,60^. Furthermore, we previously found that regions with significantly lower χ in schizophrenia corresponded to those with high oligodendrocyte gene expression^23^.

Our meta-analysis showed that the substantia nigra was the site of the greatest magnitude R2* reduction and the only significant χ reduction in psychosis. Because both measures are sensitive to all forms of iron^13^, this suggests that overall nigral iron levels are lower in psychosis. NM-MRI specifically indexes the neuromelanin-iron complex in dopamine neurones^12^. As neuromelanin is a byproduct of catecholamine metabolism^5^, elevated dopamine synthesis could drive neuromelanin accumulation, leading to the observed higher NM-MRI values in psychosis. This interpretation is supported by studies showing that nigral NM-MRI values correlate with both striatal dopamine synthesis^46^ and release capacity^42^.

A causal link between low nigral iron, elevated neuromelanin-bound iron, and striatal dopamine dysfunction in psychosis has not yet been established. Preclinical evidence shows that nigral iron deficiency leads to multiple features of striatal hyperdopaminergia observed in psychosis^61–64^. Consistent with this, our recent multimodal neuroimaging study found that lower nigral χ correlated with higher striatal dopamine synthesis capacity and greater psychotic symptom severity in people with schizophrenia^65^. These findings support a model in which low nigral iron contributes to hyperdopaminergia and subsequent neuromelanin accumulation. Because neuromelanin strongly chelates iron and does not release it under physiological conditions^5^, this sequestration would further deplete the bioavailable iron pool. Experimental studies directly examining how iron deficiency affects dopamine and neuromelanin synthesis in dopaminergic neurons are needed to test this hypothesis.

## Conclusions

This meta-analysis links psychosis with lower subcortical iron-sensitive MRI values—most prominently in the substantia nigra—alongside higher nigral NM-MRI values, which index neuromelanin-iron within dopamine neurones. Preclinical evidence indicates that nigral iron deficiency can induce striatal hyperdopaminergia, and increased dopamine turnover promotes neuromelanin synthesis, outlining a potential pathway linking low nigral iron to elevated dopamine and neuromelanin synthesis in psychosis. Examining the mechanisms underlying the interaction between these factors may reveal insights into disease pathophysiology and identify new treatment targets.

## Data Availability

Publicly available data.

## Acknowledgements and disclosures

For the purpose of open access, this paper has been published under a creative commons license (CC-BY) to any accepted author manuscript version arising from this submission. This study was funded by Medical Research Council-UK (MC_U120097115; MR/W005557/1 and MR/V013734/1), and Wellcome Trust (no. 094849/Z/10/Z) grants to Dr Howes and the National Institute for Health and Care Research (NIHR) Biomedical Research Centre at South London and Maudsley NHS Foundation Trust and King’s College London. Dr McCutcheon’s work is funded by a Wellcome Trust Clinical Research Career Development Fellowship (224625/Z/21/Z). He is also supported by the NIHR Oxford Health Biomedical Research Centre. The views expressed are those of the author(s) and not necessarily those of the NIHR or the Department of Health and Social Care.

Dr Bukala has served as a consultant for Candesic and Relation Therapeutics. Dr McCutcheon has received speaker/consultancy fees from Karuna, Janssen, Boehringer Ingelheim, and Otsuka, and co-directs a company that designs digital resources to support treatment of mental illness. Dr Howes has received investigator-initiated research funding from and/or participated in advisory/speaker meetings organized by Angellini, Autifony, Biogen, Boehringer-Ingelheim, Eli Lilly, Elysium, Heptares, Global Medical Education, Invicro, Jansenn, Karuna, Lundbeck, Merck, Neurocrine, Ontrack/Pangea, Otsuka, Sunovion, Recordati, Roche, Rovi and Viatris/Mylan. He was previously a part-time employee of Lundbeck A/v. Dr Howes has a patent for the use of dopaminergic imaging. All other authors report no biomedical financial interests or potential conflicts of interest.

