## Supplemental Materials for "Grey Matter Iron and Neuromelanin in Psychosis: A Systematic Review and Meta-Analysis of MRI Studies"

**Supplementary Figures and Tables**

Figure S1. PRISMA flow diagram for inclusion of records

Records identified from*:

Databases (n = 950)

Registers (n = 0)

**Identification of studies via databases and registers**

Records removed *before screening*:

Duplicate records removed (n = 198)

Records removed for other reasons (n = 0)

**Identification**

Records screened

(n = 752)

Records excluded**

(n = 611)

Reports sought for retrieval

(n = 141)

Reports not retrieved

(n = 0)

**Screening**

Reports excluded (n = 114):

Other neuroimaging (n = 58)

Not reporting values (n = 22)

White matter only (n = 18)

Reviews/Errata (n = 7)

Medical comorbidities (n = 4)

Not psychosis (n = 4)

Postmortem (n = 1)

Reports assessed for eligibility

(n = 141)

Studies included in review

(n = 25)

Reports of included studies

(n = 27)

**Included**

Table S1. The Newcastle-Ottawa Quality Assessment Scale for Case Control Studies.

| **Modality** | **First author & year** | **Selection**  **(/4)** | **Comparability**  **(/2)** | **Total**  **(/6)** |
| --- | --- | --- | --- | --- |
| R2* & QSM | Xu 2021 | 3 | 2 | 5 |
|  | García Saborit 2024 | 3 | 1 | 4 |
|  | Vano 2025a | 4 | 2 | 6 |
| R2* only | Cuesta 2021 | 4 | 2 | 6 |
|  | Sui 2022 | 4 | 2 | 6 |
|  | Sonnenschein 2022 | 2 | 2 | 4 |
| QSM only | Shibukawa 2024 | 3 | 2 | 5 |
|  | Ravanfar 2022 | 3 | 2 | 5 |
|  | Vano 2025b | Same study as Vano 2025a | | |
| R2 | Andreasen 1991 | 4 | 1 | 5 |
|  | Williamson 1992 | 2 | 2 | 4 |
|  | Buckley 1995 | 2 | 2 | 4 |
|  | Supprian 1997 | 2 | 2 | 4 |
|  | Pfefferbaum 1999 | 2 | 2 | 4 |
|  | Ganzetti 2015 | 2 | 2 | 4 |
| NM-MRI | Shibata 2008 | 2 | 2 | 4 |
|  | Sasaki 2010 | 2 | 2 | 4 |
|  | Watanabe 2014 | 3 | 2 | 5 |
|  | Yamashita 2016 | 3 | 2 | 5 |
|  | Cassidy 2019 | 4 | 2 | 6 |
|  | Jalles 2020 | 2 | 2 | 4 |
|  | Choi 2023 | 4 | 2 | 6 |
|  | Slifstein 2024 | 3 | 2 | 5 |
|  | Vano 2024 | Same study as Vano 2025a | | |
|  | van Hooijdonk 2023 | 4 | 2 | 6 |
|  | van der Pluijm 2024 | 4 | 2 | 6 |
|  | Wengler 2024 | 4 | 2 | 6 |

Newcastle-Ottawa Quality Assessment Scale star scores for each category and study. The exposure category was omitted as it is not applicable to imaging studies. R2*, effective transverse relaxation rate; QSM, quantitative susceptibility mapping; R2, transverse relaxation rate; NM-MRI, neuromelanin-sensitive MRI.

Figure S2. Forest plot showing the study effect sizes of the effective transverse relaxation rate (R2*) for each region of interest.

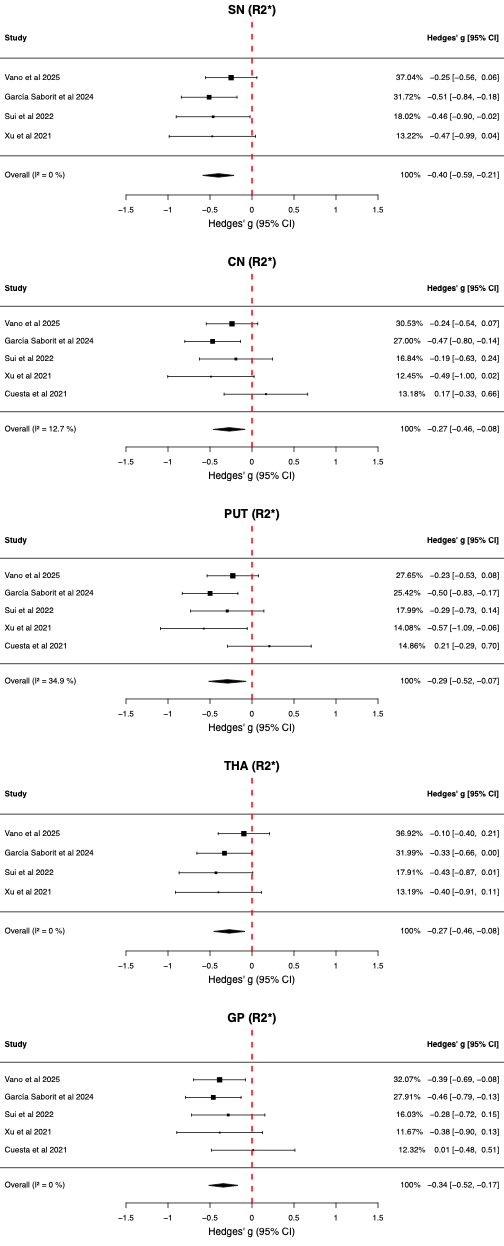

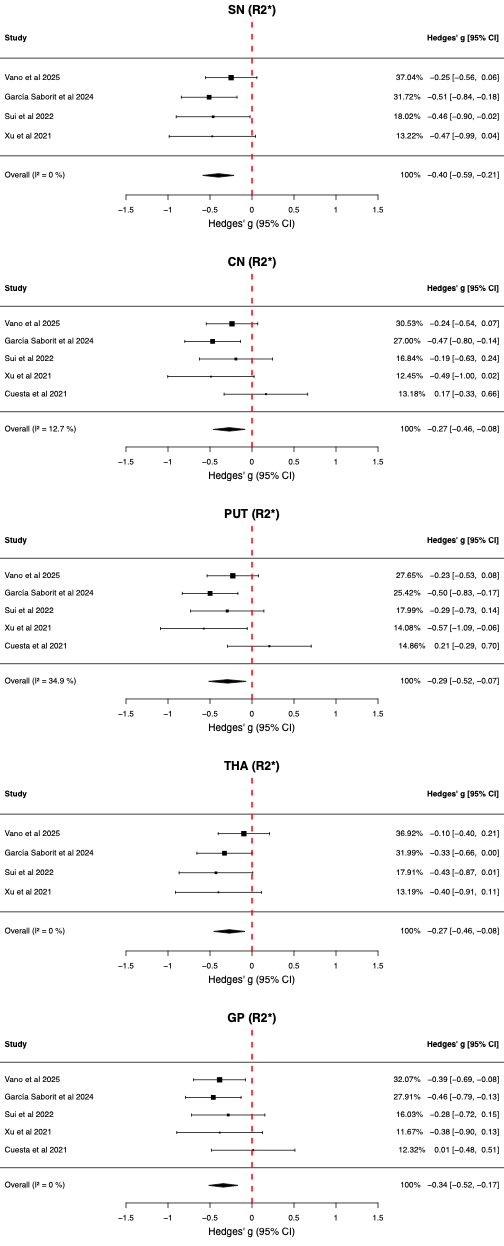

Abbreviations: CN, caudate nucleus; PUT, putamen; GP, globus pallidus; THA, thalamus; SN, substantia nigra; CI, confidence interval.

Figure S3. Forest plot showing the study effect sizes of the magnetic susceptibility (χ), calculated by quantitative susceptibility mapping (QSM), for each region of interest.

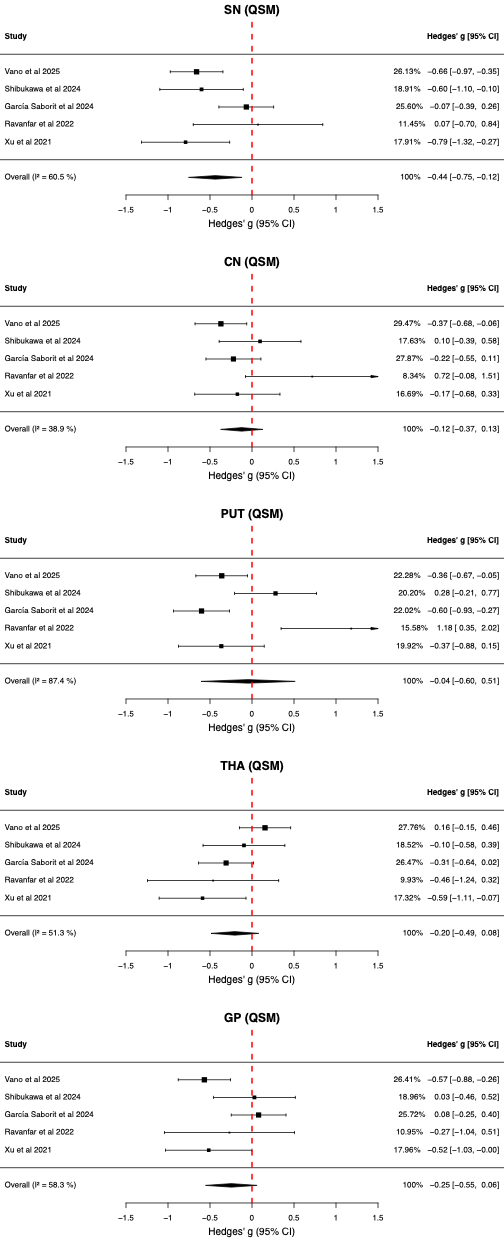

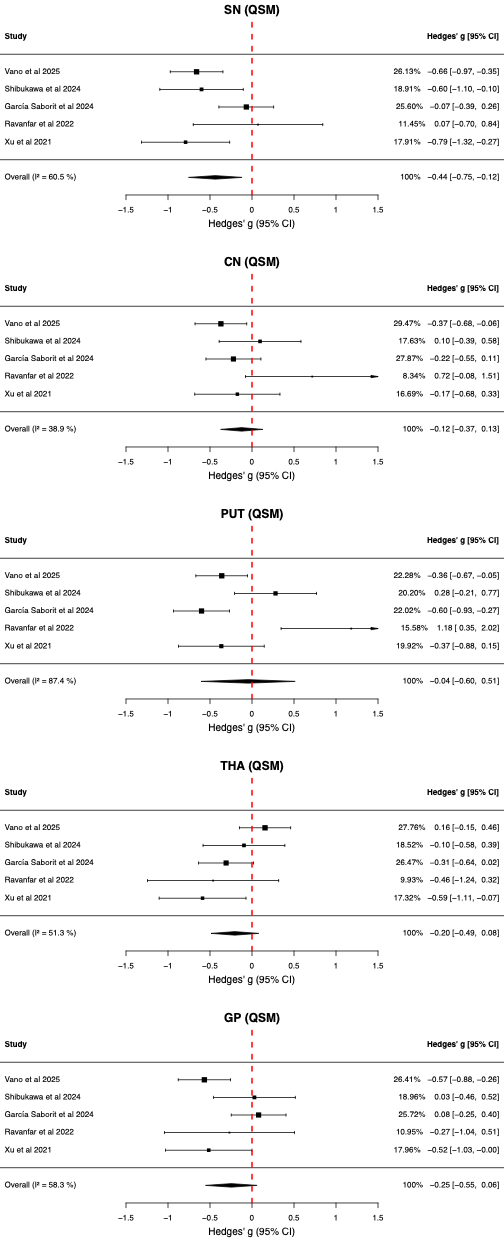

Abbreviations: CN, caudate nucleus; PUT, putamen; GP, globus pallidus; THA, thalamus; SN, substantia nigra; CI, confidence interval.

Figure S4. Forest plot showing the study effect sizes of the transverse relaxation rate (R2) for each region of interest.

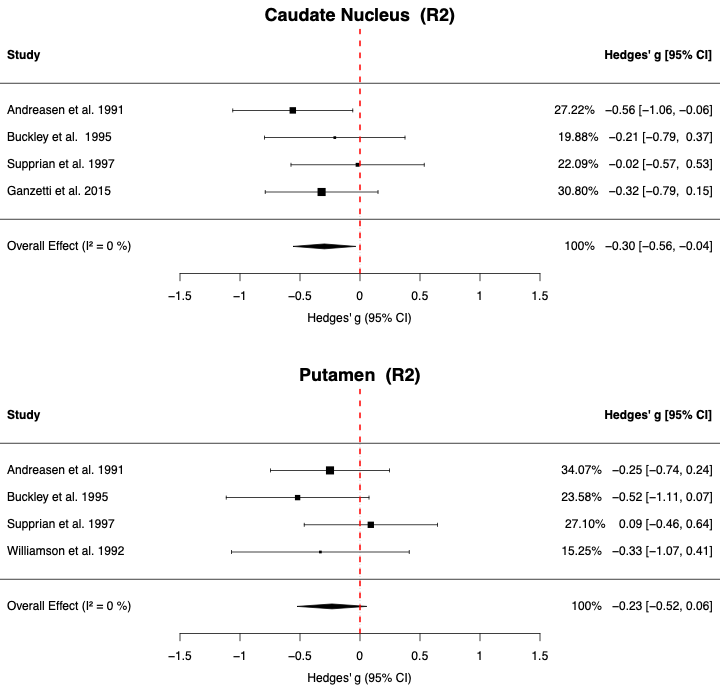

Abbreviations: CN, caudate nucleus; PUT, putamen; CI, confidence interval.

Figure S5. Forest plot showing the study effect sizes of the neuromelanin-sensitive MRI (NM-MRI) values for the substantia nigra.

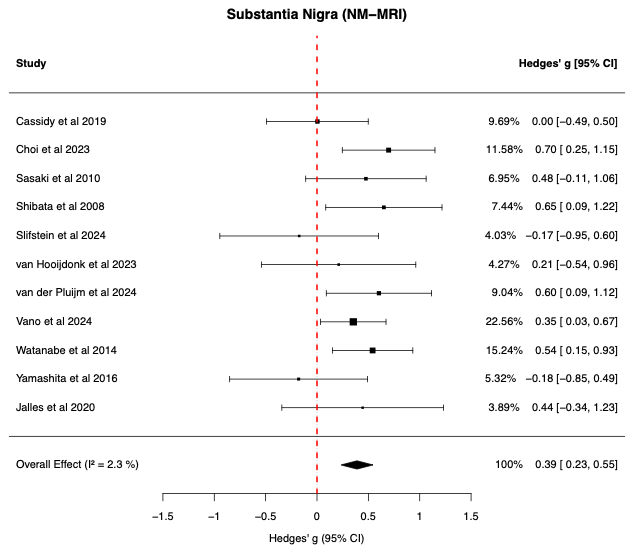

Abbreviations: CI, confidence interval.

Figure S6. Graph showing the group-average magnetic susceptibility (χ) calculated by quantitative susceptibility mapping (QSM) for each study.

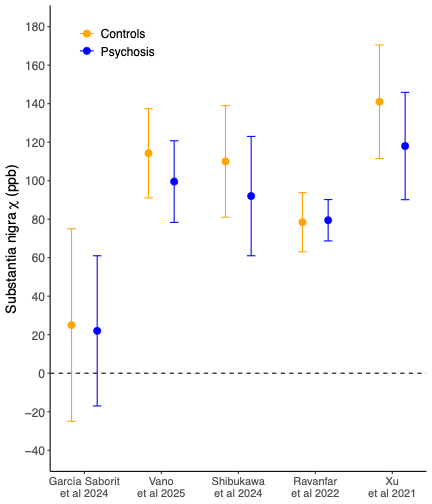

Group means ± standard deviation are shown. Abbreviations: ppb, parts per billion.

Table S2. Results from the leave-one-out (LOO) and variability analyses

| MRI modality | ROI | LOO minimum | LOO Maximum | Log SD ratio | Log SD 95% CI | Log SD ratio p-value |
| --- | --- | --- | --- | --- | --- | --- |
| R2* | SN | -0.49 | -0.35 | -0.05 | [-0.18, 0.08] | 0.490 |
|  | CN | -0.34 | -0.20 | 0.11 | [-0.09, 0.31] | 0.290 |
|  | PUT | -0.37 | -0.22 | -0.03 | [-0.25, 0.20] | 0.811 |
|  | THA | -0.37 | -0.24 | 0.09 | [-0.10, 0.28] | 0.361 |
|  | GP | -0.39 | -0.29 | -0.04 | [-0.16, 0.08] | 0.506 |
| QSM | SN | -0.67 | -0.53 | -0.07 | [-0.23, 0.08] | 0.356 |
|  | CN | -0.22 | -0.02 | 0.07 | [-0.06, 0.20] | 0.292 |
|  | PUT | -0.29 | 0.12 | 0.05 | [-0.29, 0.38] | 0.787 |
|  | THA | -0.33 | -0.12 | -0.08 | [-0.21, 0.05] | 0.237 |
|  | GP | -0.37 | -0.11 | -0.06 | [-0.23, 0.12] | 0.527 |
| R2 | CN | -0.37 | -0.20 | 0.08 | [-0.14, 0.30] | 0.489 |
|  | PUT | -0.35 | -0.14 | 0.30 | [0.09, 0.50] | 0.004 |
| NM-MRI | SN | 0.35 | 0.43 | 0.11 | [-0.12, 0.33] | 0.345 |

Abbreviations: SD, standard deviation; CI, confidence interval; R2*, effective transverse relaxation rate; QSM, quantitative susceptibility mapping; R2, transverse relaxation rate; NM-MRI, neuromelanin-sensitive MRI; SN, substantia nigra; CN, caudate nucleus; PUT, putamen; THA, thalamus; GP, globus pallidus.

Figure S7. The correlation between substantia nigra neuromelanin-sensitive MRI (NM-MRI) effect size and chlorpromazine daily equivalent dose.

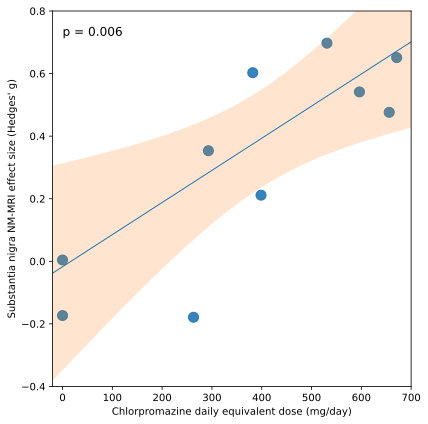

Figure S8. Summary plot showing effect sizes and heterogeneity estimates in the caudate nucleus and putamen for each iron-sensitive MRI modality when studies were separated by first-episode psychosis (FEP) and chronic psychosis.

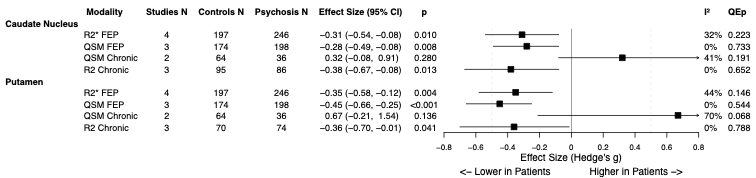

Effective transverse relaxation rate (R2*) and quantitative susceptibility mapping (QSM)-derived magnetic susceptibility (χ) were significantly lower for the caudate nucleus and putamen in the FEP groups compared to controls, with no significant heterogeneity. While no case-control significant differences were seen in the chronic cohorts for these measures, the tissue transverse relaxation rate (R2) was significantly lower for both regions in chronic illness.

Figure S9. Funnel plot assessing publication bias of neuromelanin-sensitive MRI (NM-MRI) substantia nigra (SN) studies.

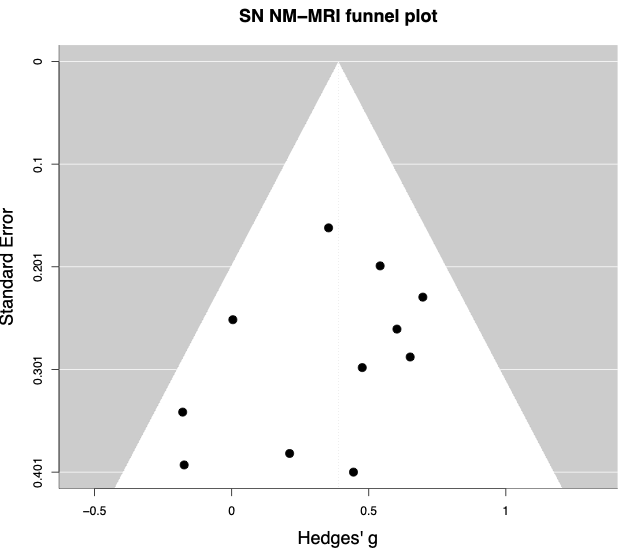

**Supplementary Methods**

Full search strategy

Embase search:

susceptibility weighted imaging/ or t2 weighted imaging/ or exp nuclear magnetic resonance imaging/ and iron.ti, ab, kw. or exp nuclear magnetic resonance imaging/ and neuromelanin.ti, ab, kw. or (QSM or "quantitative susceptibility mapping").ab, ti, kw. or exp nuclear magnetic resonance imaging/ and R2.ti, ab, kw. or exp nuclear magnetic resonance imaging/ and R²*.ti, ab, kw. or exp nuclear magnetic resonance imaging/ and T2*.ti, ab, kw. or exp nuclear magnetic resonance imaging/ and T2.ti, ab, kw. And ("psychosis" or "schizo*" or "psychotic" or "bipolar disorder" or "manic" or "bipo*").ti, ab, kw. and article.pt.

PsycINFO search:

magnetic resonance imaging/ and ("QSM" or "quantitative susceptibility mapping" or "iron" or "iron*" or "susceptibility weighted imaging" or "SWI" or "relaxometry" or "gradient echo" or "gradient recall echo" or "GRE" or "T2" or "R2" or "R2*" or "T2*" or "neuromelanin").mp. AND ("psychosis" or "schizo*" or "psychotic" or "bipolar disorder" or "manic" or "bipo*").mp.

PubMed search:

"Magnetic Resonance Imaging"[Mesh] and ("QSM" or "quantitative susceptibility mapping" or "iron" or "iron*" or "susceptibility weighted imaging" or "SWI" or "relaxometry" or "gradient echo" or "gradient recall echo" or "GRE" or "T2" or "R2" or "R2*" or "T2*" or "neuromelanin") and "Schizophrenia Spectrum and Other Psychotic Disorders"[Mesh]

Selection of studies

Identified studies from all databases were combined, and duplicates were removed. Each of the remaining studies was allocated to two investigators (either BB, RC, or LJV) who independently screened the titles and abstracts against our eligibility criteria. For all studies marked for inclusion by at least one investigator, full-text records were obtained. Both allocated investigators reviewed these records to decide on inclusion for data extraction. If a consensus was not reached, a third reviewer (RAM) made the ultimate decision.

Data collection and analysis

Data collection and management

Both allocated investigators independently collected data from each selected paper. Any disagreements were discussed and resolved by the third author (RAM).

We converted relevant metrics to enable cross-study comparisons. The Brief Psychiatric Rating Scale (BPRS) total scores were converted to Positive and Negative Symptom Score (PANSS) totals using an equipercentile linking method^1^. The Scale for the Assessment of Positive Symptoms (SAPS) and Scale for the Assessment of Negative Symptoms (SANS) were converted to PANSS positive and negative scores, respectively^2^. Additionally, the olanzapine equivalence doses were converted to a CPZE doses^3^.

**Supplementary Analyses**

Narrative review

ROIs analysed in fewer than 4 studies

- **Globus pallidus and thalamus:** Globus pallidus was examined in two R2 studies ^4,5^, and thalamus in one R2 study ^6^. No significant difference between patients with chronic schizophrenia and matched controls.
- **Red nucleus:** examined in two R2* ^7,8^ and three QSM studies ^7–9^. Only one QSM study identified a significant case–control difference (controls N = 30; first-episode schizophrenia N = 30; g = −0.79, 95% CI [−1.32, −0.27], p = 0.003) ^8^.
- **Nucleus accumbens:** examined in two R2* ^7,10^ and three QSM studies ^7,9,11^. Only one QSM study identified a significant difference (controls N = 50; chronic psychosis N = 24; g = 0.50, 95% CI [0.01, 1], p = 0.045) ^9^.
- **Amygdala:** examined in one QSM ^9^ and two R2 studies ^6,12^. No significant difference between patients with chronic schizophrenia and matched controls.
- **Hippocampus:** examined in two QSM ^9,11^ and one R2 study ^13^. Only the R2 study reported a significant difference (controls N = 10; chronic psychosis N = 24; g = −0.75, 95% CI [−1.51, −0.01], p = 0.049) ^13^.

**Other cortical regions.** Only examined using R2 in chronic schizophrenia.

- **Whole cortex:** examined by two studies ^5,14^; one reported a significant reduction (controls N = 10; chronic psychosis N = 10; g = −1.36, 95% CI [−2.34, −0.38], p = 0.007) ^14^.
- **Frontal cortex:** examined by two studies ^6,12^; one reported a significant reduction (controls N = 38; psychosis N = 27; g = −0.67, 95% CI [−1.18, −0.17], p = 0.009) ^12^.
- **Dorsolateral cortex:** examined by two studies ^12,13^; one reported a significant reduction (controls N = 38; psychosis N = 27; g = −0.71, 95% CI [−1.21, −0.20], p = 0.006) ^12^.
- **Cingulate cortex:** examined by one study ^12^; no significant difference.
- **Temporal cortex:** examined by two studies ^12,14^; no significant difference.

**NM-MRI.** Most studies examined the dopaminergic midbrain as a whole structure, often calling this the substantia nigra. Below are the studies that examined components separately, and those that examined the noradrenergic midbrain.

- **Substantia nigra pars compacta and pars reticulata:** one study separately examined these structures and found no significant difference for either ^15^.
- **Ventral tegmental area:** examined by two studies ^15,16^, with one study finding a significant difference (controls N = 22; psychosis N = 14; g = −1.44, 95% CI [−2.19, −0.69], p < 0.001) ^16^.
- **Locus coeruleus**: Examined by three studies ^17–19^; no significant difference.

Voxelwise analysis

Sonnenschein et al. identified voxels in the right ventral lateral thalamus where R2* was higher for 72 patients with chronic schizophrenia than for 74 healthy controls ^20^. Our recent QSM paper also found a cluster predominantly in this right ventral lateral thalamus where χ was higher in patients with schizophrenia than controls. We also associated schizophrenia with higher χ for a cluster in the right nucleus accumbens and lower χ for seven clusters found bilaterally in the substantia nigra, putamen, globus pallidus, and the left caudate nucleus ^7^.

In the same cohort our NM-MRI analysis highlighted voxels in the bilateral ventromedial substantia nigra where NM-MRI signal was significantly greater in patients with schizophrenia than controls ^21^. Cassidy et al. identified voxels prominently in the ventral substantia nigra where increased NM-MRI signal correlated with greater psychotic symptom severity ^22^. This region was also identified as important by van der Pluijm et al., who associated higher NM-MRI signal here with positive treatment response to antipsychotic medications in first-episode psychosis ^23^.

Psychosis subgroup analyses

Watanabe et al. examined the effect of age on NM-MRI group differences ^19^. Greater NM-MRI signal elevation was observed for patient with psychosis in those < 30 years old (controls N = 29; psychosis N = 24; g = 0.81, 95% CI [0.24, 1.37], p = 0.005), but not in those ≥ 30 years old (controls N = 23; psychosis N = 28; g = 0.25, 95% CI [-0.31, 0.80], p = 0.377).

Cassidy et al. separated the psychosis group by symptom severity ^22^. They found that compared to controls, NM-MRI signal was significantly elevated in the severely psychotic group (controls N = 30; psychosis N = 9; g = 0.92, 95% CI [0.15, 1.70], p = 0.020), but not the mild psychosis group (controls N = 30; psychosis N = 24; g = -0.19, 95% CI [-0.73, 0.35], p = 0.490).

Only one study has directly compared the case-control effect for different psychotic diagnostic categories ^24^. Sui et al. examined synthetic R2* in the caudate nucleus, putamen, globus pallidus, thalamus, and ventral diencephalon (grouped with the substantia nigra in our analyses) in 35 healthy controls, 15 patients with schizophrenia, 17 with schizoaffective disorder, and 17 with bipolar disorder. Using ANCOVA, the authors found that patients with schizophrenia had significantly lower R2* than controls in all regions except the caudate nucleus (80% of ROIs). Patients with schizoaffective disorder showed significant reductions in 50% of the ROIs, generally with smaller effect sizes than in schizophrenia, while no significant differences were observed in bipolar disorder.

van der Pluijm et al. used NM-MRI to compare patients with schizophrenia who responded to antipsychotic treatment and those with treatment-resistant schizophrenia with matched healthy controls ^23^. Compared to controls, the treatment responders had significantly higher NM-MRI values (controls N = 20; psychosis N = 47; g = 0.61, 95% CI [0.07, 1.14], p = 0.027), while treatment-resistant patients showed no significant differences (controls N = 20; psychosis N = 15; g = -0.06, 95% CI [-0.73, 0.61], p = 0.861).
